# RENOVO-NF1 accurately predicts NF1 missense variant pathogenicity

**DOI:** 10.1101/2025.01.06.25320039

**Authors:** Emanuele Bonetti, Serena Pellegatta, Nayma Rosati, Marica Eoli, Luca Mazzarella

## Abstract

The identification of a pathogenic variant in the NF1 gene is an important step in the diagnosis of the tumor-predisposing and developmental syndrome neurofibromatosis, and is increasingly important in the characterization of sporadic tumors, in which NF1 loss identifies specific biologic subtypes. However, NF1 variant interpretation is complicated by multiple factors including allelic heterogeneity, sequence homology, and the lack of functional assays to confirm loss of function. Computational tools able to predict variant pathogenicity may represent an ideal complement to the lengthy process of clinical validation, often impossible within an adequate time frame.

Here, we present RENOVO-NF1, an evolution of our previously published tool RENOVO algorithm, optimized for the interpretation of NF1 variants. RENOVO-NF1 has an accuracy of 98.6% on (likely) pathogenic/(likely) benign (P/LP/B/LB) variants, and importantly shows high accuracy on P/LP/B/LB variants that were initially classified as of unknown significance (VUS), in particular missense. Based on this, we predict reclassification for 79% of the variants currently classified as missense VUS in ClinVar. Since missense VUS are the most represented and problematic variant class, RENOVO-NF1 may significantly aid in diagnostic challenges in neurofibromatosis and in precision oncology for putatively NF1-mutated patients.

## Introduction

Neurofibromatosis is a complex developmental syndrome characterized by dermatological, neurological, skeletal and cardiovascular defects and a predisposition to multiple benign and malignant tumors. Neurofibromatosis is caused by loss of function in neurofibromin, a large protein coded by the NF1 locus in 17q11.2. Although the diagnosis of neurofibromatosis is primarily based on clinical findings, the detection of pathogenic variants in NF1 was recently introduced among the revised diagnostic criteria (Legius et al., 2021). This is particularly useful for the early diagnosis in oligosymptomatic children without a family history of the disease, who would not otherwise meet the strict clinical criteria (Kehrer-Sawatzki and Cooper, 2022). The interpretation of NF1 variants is complicated by several factors: locus size, high sequence homology, significant allelic heterogeneity with several mutations arising de novo in the absence of a suggestive family history (thus preventing segregation analysis), the high mutation frequency (one of the highest in the human genome, (Huson et al., 1989; Clementi et al., 1990) and the still incompletely understood biology of the coded protein, neurofibromin, which prevented the development of confirmatory functional assays.

In 2020 we developed RENOVO, a random forest-based algorithm for variant interpretation that achieved superior accuracy over existing algorithms (Favalli et al., 2021). RENOVO provides a Pathogenicity Likelihood Score (PLS) which indicates the percentage of decision trees agreeing on the interpretation. The PLS, its associated probability and optimal cutoff points were originally computed under a generalized framework for variant interpretation of any gene. However, we showed that for some diseases (e.g. dilated cardiomyopathies) or genes (e.g. SCN5A), modifying the cutoff points enhances accuracy. An original aspect of RENOVO development was the construction of the datasets through an approach of “database archaeology”. By comparing versions of ClinVar over the years, we defined a set of “stable” variants that maintained the same pathogenic/likely pathogenic/benign/likely benign (P/LP/B/LB) classification over time, and a set of “unstable” variants that were initially classified as Variants of Unknown Significance (VUS) but were subsequently reclassified as P/LP or B/LB. Stable and unstable variants constituted the training and test set, respectively. This approach allows to retrospectively measure algorithm accuracy on classification prediction, the scenario in which variant interpretation algorithms are most useful. Recently, RENOVO predictions were further assessed prospectively, on a third validation dataset with variants that were classified as VUS in ClinVar and scored by RENOVO in 2020, but were subsequently reclassified as P/B over the ensuing 4 years. RENOVO maintained high accuracy (82%) on the prospective validation, with peaks approaching 100% for specific genes and variant types (Bonetti et al., 2024).

Here, after constructing NF1-specific datasets nested within the training, test and validation datasets, we redefined optimal parameters for RENOVO application to NF1 variants, and demonstrated the accuracy of RENOVO-NF1 on VUS predictions. Finally, we propose a reclassification for 2978 missense variants currently classified as VUS.

## Results

Figure 1 presents an analysis of the current state of NF1 variants in ClinVar. As of July 2024, 13460 NF1 variants are listed; most (5082, 37.7%) are classified as VUS. Missense variants are the most represented type (5198, 38.6%); for missense, the percentage of VUS is significantly higher (4412, 84.8%), highlighting the need to support variant interpretation with informatic tools for this variant class.

**Figure 1.**
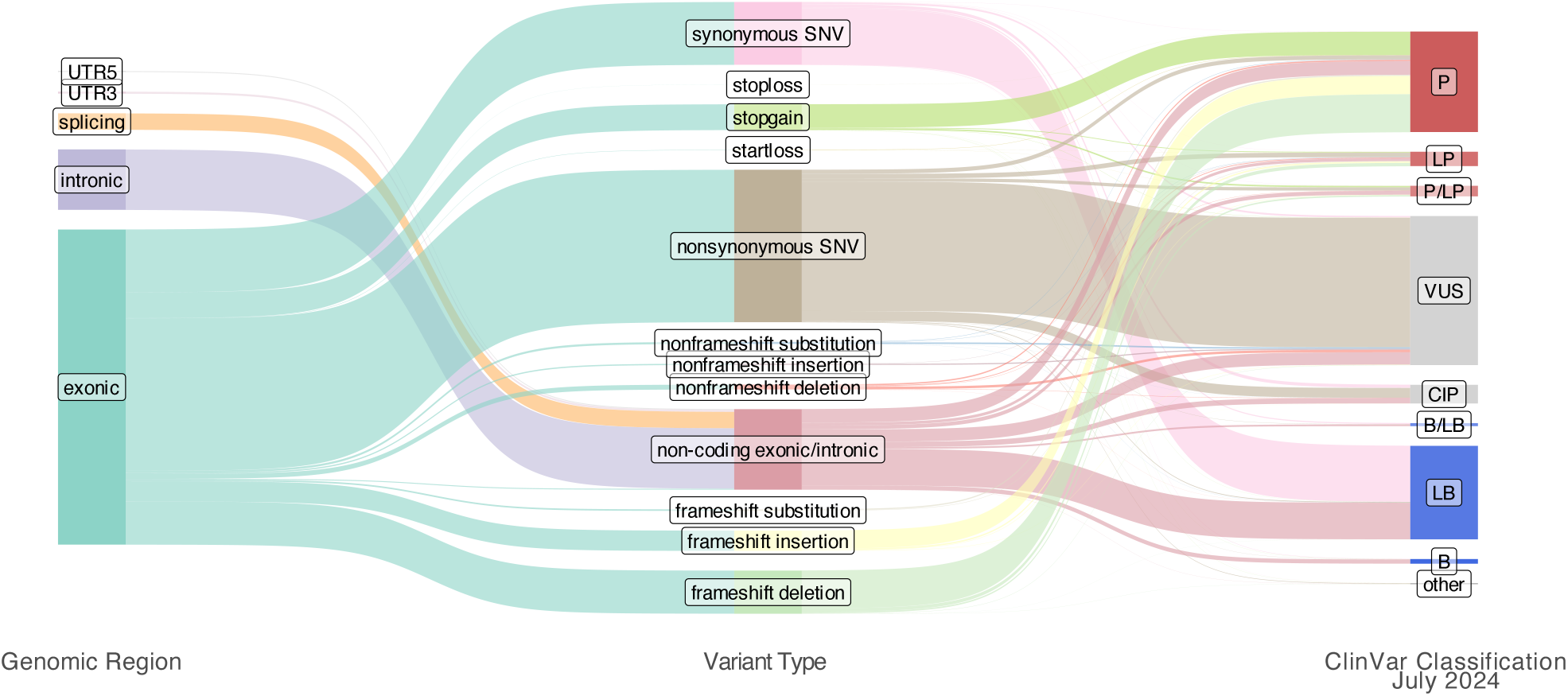
Sankey diagram representing the state of NF1 variants in ClinVar as of July 2024

To optimize RENOVO for NF1 assessment, we calculated NF1-specific optimal PLS cutoff points on the 2020 training set (n=2375, 1171 P/LP, 1204 B/LB), maximizing the “accuracy” metric. We identified the cutoffs at 0.6465 for P/LP and 0.4278 at B/LB (supplementary figure 1A-B). Variants with a PLS higher than 0.6465 were defined as RENOVO-NF1 pathogenic (RNF1-P); variants with a PLS lower than 0.4278 were defined as RENOVO-NF1 benign (RNF1-B); variants with a PLS intermediate between the 2 cutoff points were considered as RENOVO-NF1 uncertain (RNF1-U). With these parameters, accuracy in stable variants in the training set was 98.6%.

We then classified NF1 variants in the 2020 test set (n= 57, of which 36 P/LP, 21 B/LB) and in the 2024 validation set, which includes NF1 variants that were classified as VUS in 2020 and were reclassified as P/LP/B/LB in 2024 (n=100, of which 58 P/LP and 42 B/LB).

On the 2020 test set, accuracy was 96.5%, with only 2/57 inaccurate or uncertain predictions (figure 2A).

**Figure 2.**
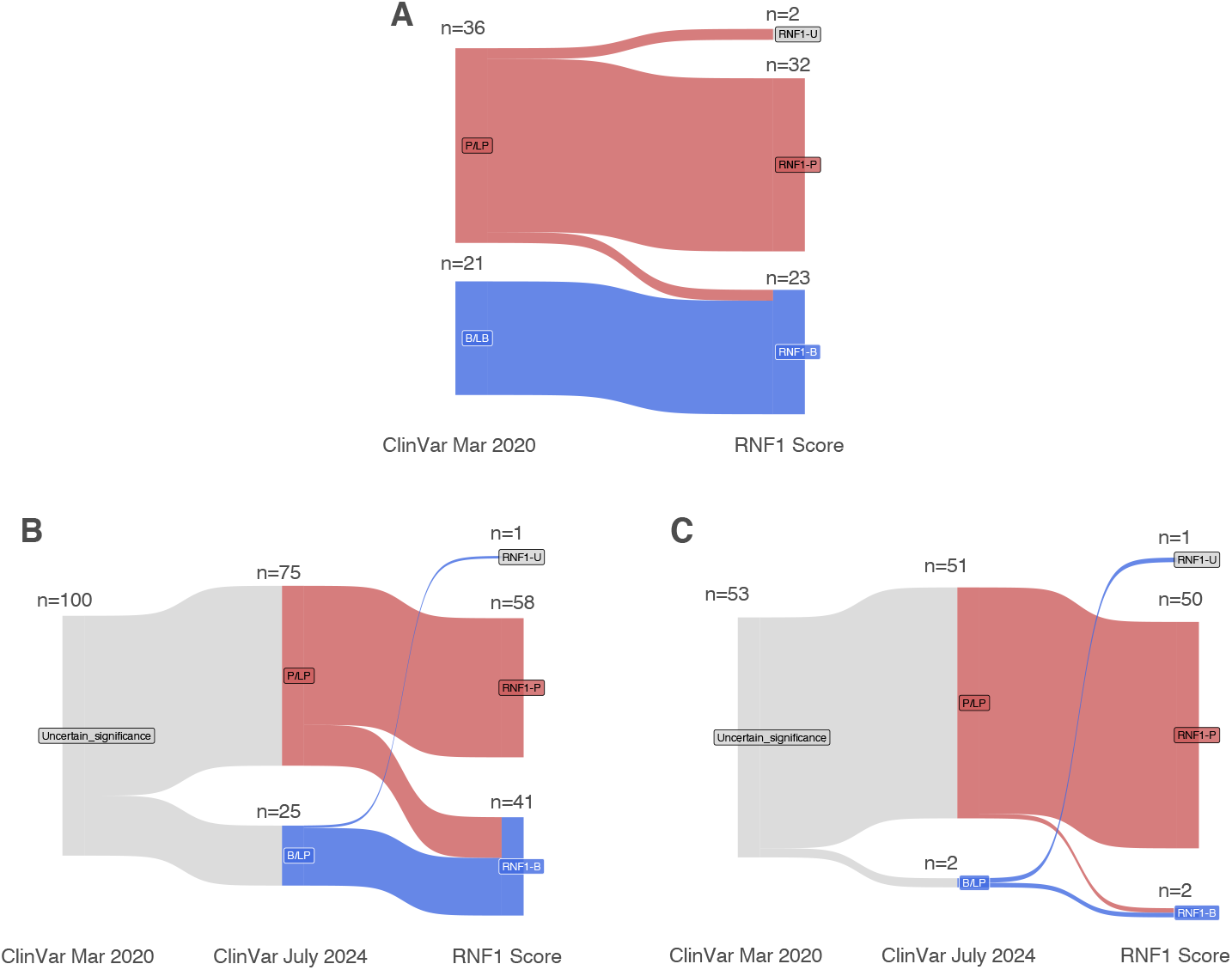
RENOVO performance on test and validation sets A) test set of 57 variants classified as VUS prior to 2020 and subsequently reclassified in P/LP (n=36) or B/LB (n=21) in 2020 B) validation set of 100 variants classified as VUS in 2020 and subsequently reclassified in P/LP (n=58) or B/LB (n=42) in 2024 C) missense variants (n=57) in the validation set

On the more challenging 2024 validation set, accuracy remained high at 82% (figure 2B). Performance metrics were conditioned by variant class. Although accuracy was suboptimal for intronic variants, it achieved 96.2% for the most represented class (nonsynonymous SNVs, n=53) (figure 2C).

Finally, we generated RENOVO-based classification of the 4412 variants listed as missense VUS in ClinVar as of July 2024. RENOVO classified 2270 (62.8%) variants as RNF1-P, 708 (16%) as RNF1-B and 934 (21.2%) RNF1-U, proposing a clinically meaningful reclassification for 79% of the NF1 missense VUSs (figure 3 and supplementary table 1).

**Figure 3.**
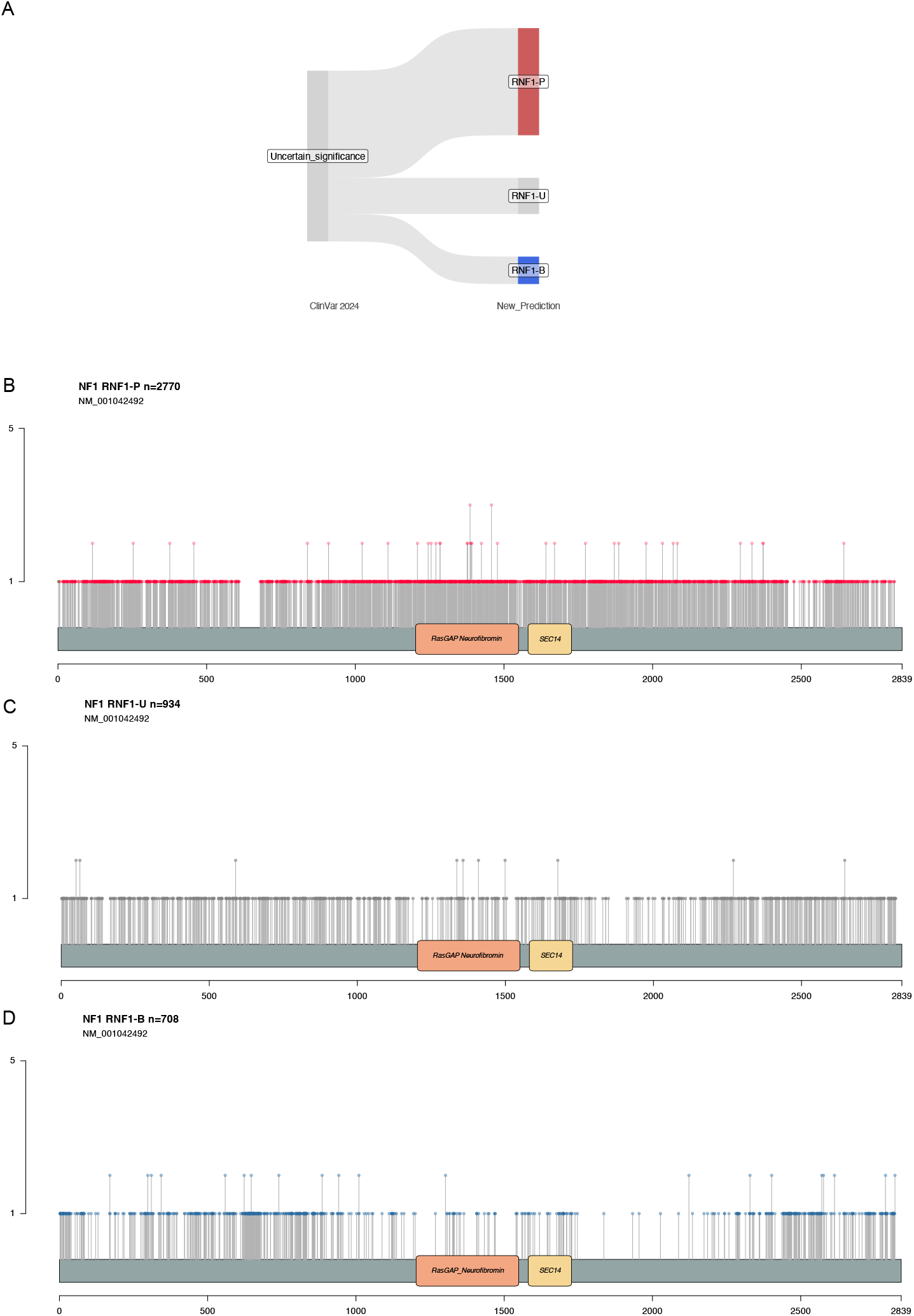
Proposed reclassification for 2270 NF1 missense variants currently classified as VUS. A) Sankey diagram of the proposed reclassification B-D) Distribution of reclassified missense variants along the NF1 gene body: RNF1-P (B), RNF1-B (C), RNF1-U (D)

## Discussion

We show here that RENOVO-NF1, an enhanced version of our algorithm RENOVO optimized for NF1, achieves high accuracy for variants for which the evidence at the moment of identification is insufficient to classify them as P/LP/B/LB. This was assessed both retrospectively, on unstable variants at the moment of RENOVO training (the 2020 test set) and confirmed prospectively, on variants reclassified between 2020 and 2024. RENOVO performance was particularly high for missense variants, which are at the same time the most common and one of the most difficult to interpret variant classes in NF1.

Performance was poorer on non-missense variants, in particular intronic variants. This was expected, since many of the key features for RENOVO interpretation, such as functional impact, are not available for non-missense variants.

The interpretation of genomic variants is a key step in the clinical assessment of hereditary diseases such as neurofibromatosis. The advent of Next Generation Sequencing has greatly facilitated the identification of novel variants, but their interpretation remains highly challenging and limits the utility of Next generation Sequencing (NGS) in clinical practice. As we recently showed, VUS reporting grows exponentially but their interpretation is significantly slower, with approximately one reclassified variant every ∼30 new VUS discovered over time (Bonetti et al., 2024). The process of variant interpretation according to ACMG guidelines remains laborious and requires data, such as family segregation and functional assays, that are often unavailable in many cases of neurofibromatosis. Thus, computational methods are increasingly advocated to facilitate the prioritization of variant to be subjected to the rigorous process of ACMG validation (Bromberg and Radivojac, 2022).

In conclusion, RENOVO-NF1 successfully identifies NF1 missense variants that should be prioritized for rigorous ACMG validation. RENOVO may be particularly useful in the many cases with de novo-arising mutations in which early diagnosis is prevented by the lack of familial segregation.

## Methods

### Data availability

All data produced in the present work are contained in the manuscript

### Databases

ClinVar data for NF1 variants were extracted from the ClinVar version 2024-07-03 available at: https://ftp.ncbi.nlm.nih.gov/pub/clinvar/vcf_GRCh37/archive_2.0/2024/clinvar_20240307.v cf.gz NF1-training, -test-and -validation datasets were derived from the training, test and validation databases described in the RENOVO repository at https://github.com/mazzalab-ieo/renovo by filtering for mutations with a genomic position overlapping the NF1 locus (chr17:29,421,995-29,704,693, ref. build hg19)

### Cutpoint optimization

All analyses were conducted in R version 4.3.1.

For cutpoint optimization, we used cutpointR version 1.1.2, maximizing the “accuracy” metric for: i) class “1” variants, corresponding to ClinVar “Pathogenic”, “Pathogenic/Likely_pathogenic” and “Likely_pathogenic” and ii) class “0” variants, corresponding to “Benign”, “Benign/Likely_bening” and “Likely_bening”.

To enhance readability, we combined the ClinVar pathogenic and benign classes into two main categories: “Pathogenic”, “Pathogenic/Likely_pathogenic”, and “Likely_pathogenic” were labeled as “P”, while “Benign”, “Benign/Likely_benign”, and “Likely_benign” were labeled as “B”.

Variants with a PLS higher than or equal to 0.6465 were defined as RENOVO-NF1 pathogenic (RNF1-P); variants with a PLS lower than 0.4278 were defined as RENOVO-NF1 benign (RNF1-B); variants with a PLS intermediate between the 2 cutoff points were considered as RENOVO-NF1 uncertain (RNF1-U). We considered the following definitions for true/false positive/negative:

-a RNF1-P variant that was classified as P/LP in ClinVar 2024 → true positive

-a RNF1-B variant that was classified as B/LB in ClinVar 2024 → true negative

-a RNF1-P variant that was classified as B/LB in ClinVar 2024 → false positive

-a RNF1-B variant that was classified as P/LP in ClinVar 2024 → false negative

-a RNF1-U variant that was classified as P/LP → false negative

-a RNF1-U variant that was classified as B/LB → false negative

