## Supplementary figures and images for "RENOVO-NF1 accurately predicts NF1 missense variant pathogenicity"

### supplementary figure 1

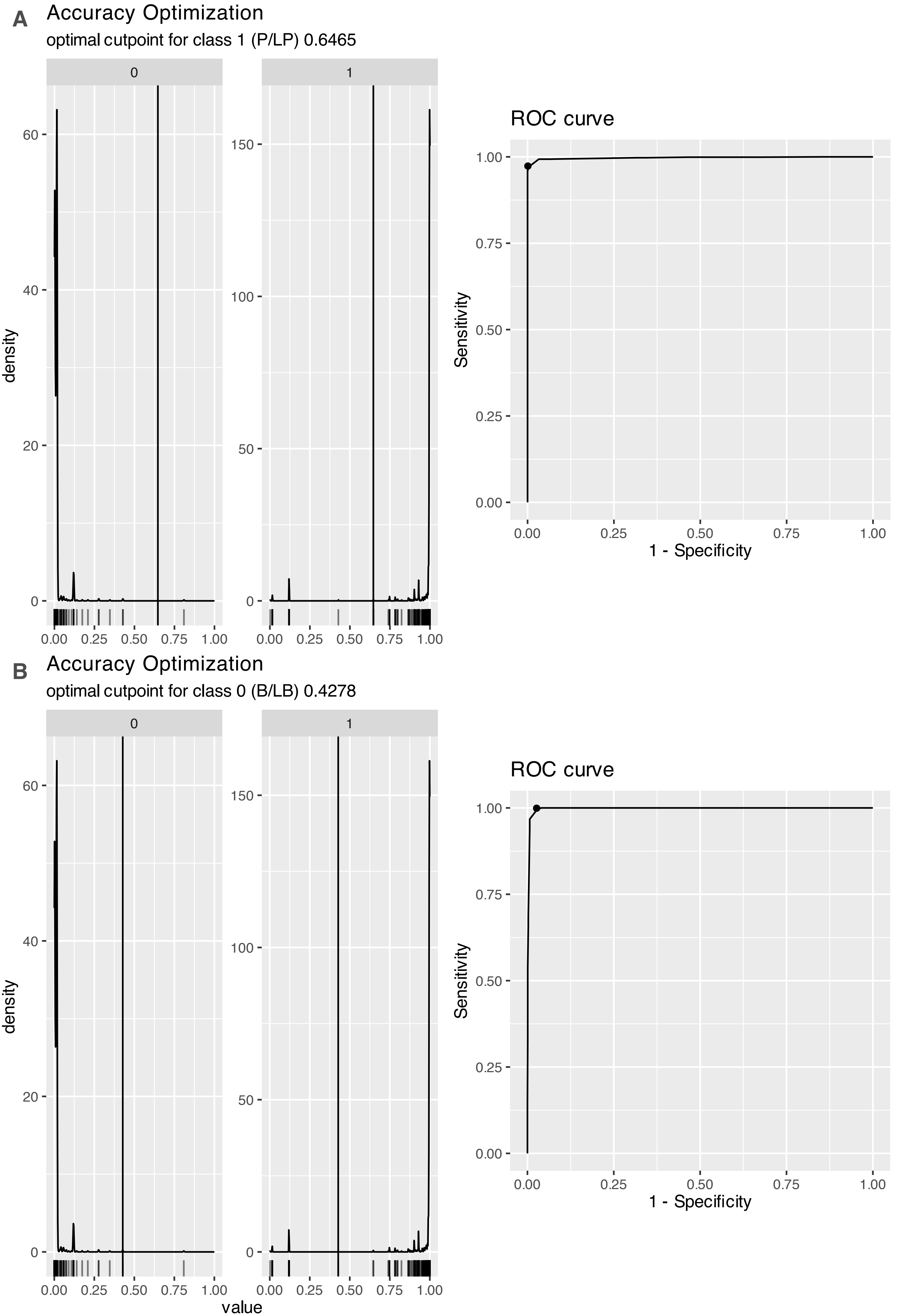
